# A Randomized Trial of a Wearable UV dosimeter for Skin Cancer Prevention

**DOI:** 10.1101/2021.11.29.21267005

**Authors:** Emmanuel LP Dumont, Peter D Kaplan, Catherine Do, Shayak Banerjee, Melissa Barrer, Khaled Ezzedine, Jonathan H Zippin, George I Varghese

## Abstract

**Background:** Non-melanoma skin cancer (NMSC) is the most prevalent cancer in the United States. Despite guidelines on ultraviolet (UV) avoidance, it remains difficult for people to assess their exposure, as UV is invisible and the onset of UV-induced symptoms is delayed.

**Methods:** In a prospective randomized trial, ninety-seven elderly patients with a history of actinic keratoses (AK) were enrolled and followed over six months. Fifty patients received UV counseling by a dermatologist and a wearable UV dosimeter that provided real-time and cumulative UV exposure. Forty-seven patients solely received UV counseling by a dermatologist.

**Results:** Over 75% of participants recorded UV exposure at least once a week during the summer. After 6 months of intervention, when comparing the device group to the control group, we observed a non-significant 20% lower ratio of incidence rates of AKs (95% CI = [-41%, 55%], p-value = 0.44) and a significant 95% lower ratio of incidence rates of NMSCs (95% CI = [33%, 99.6%], p-value = 0.024). Self-reported surveys demonstrated that the control group’s score in self-perceived ability to participate in social activities significantly increased by 1.2 (p-value = 0.04), while in the device group, this score non-significantly decreased by 0.9 (p-value = 0.1). Finally, we did not observe changes, or between-group differences, in self-reported anxiety and depression surveys.

**Conclusion:** This pilot clinical trial has a short duration and a small sample size. However, device adherence and quality of life questionnaires suggest a smartphone-connected wearable UV dosimeter is well accepted by an elderly population. This trial also indicates that a wearable UV dosimeter may be an effective behavioral change tool to reduce NMSC incidence in an elderly population with a prior history of AKs.

**Trial Registration:** The trial was registered on clinicaltrials.gov under the identification NCT03315286.

## BACKGROUND

Skin cancer is the most common cancer in the United States.[1] Genetic, phenotypic, and environmental factors, specifically ultraviolet radiation (UV), are considered the most significant contributing factors to the development of skin cancer.[2] Recent controversies on the effectiveness[3] and safety[4] of sunscreens have created a need for safer strategies to help manage UV exposure. UV dosimeters provide a data-driven solution for assessing and communicating the real-time risk of UV. In this prospective, randomized clinical trial, we evaluated the clinical efficacy of a UV dosimeter and mobile app on pre- and cancerous lesions against the standard of care over six months overlapping a summer in an elderly patient population disposed to developing skin cancer.

## METHODS

### Study design

This prospective, randomized, observer-blinded, controlled clinical trial enrolled elderly patients with a history of actinic keratoses at a single site in New York City, NY. The trial was conducted under the oversight of the Institutional Review Board (IRB) of Weill Cornell Medicine and the National Cancer Institute (NCI). It adhered to applicable governmental regulations. The IRB and the NCI approved the protocol and the consent forms. As a requirement of contract HHSN261201700005c with NCI, the protocol and all amendments were submitted and approved by the program officer. All participants provided written informed consent before enrollment. The sponsor, YouV Labs, Inc., and the trial’s principal investigator (GV) were responsible for the overall trial design, site selection, monitoring, and data analysis. Investigators were responsible for data collection, recruitment, and treatment. The authors vouch for the accuracy and completeness of the data and the fidelity of the trial to the protocol. The trial was registered on clinicaltrials.gov under the identification NCT03315286 on October 20, 2017.

### Participants, randomization, and data blinding

Eligible participants were 18 years of age or older with a history of actinic keratosis (AKs, one diagnosis in the 12 months before enrollment or 5 AKs in the five years before enrollment). Patients having received UV therapy in the past six months or field therapy for the treatment of actinic keratosis in the past three months were excluded. Participants were assigned using randomly-generated blocks of four, stratified by skin type, to receive a wearable UV dosimeter and standard-of-care UV education or solely standard-of-care UV education (avoid going outside between 10 am and 4 pm, apply sunscreen with SPF 30-50 and re-apply every two hours including when coming out of the water, wear sun protective clothing, such as a hat.). The UV education was provided in person by the study dermatologist at the end of all three clinical visits. We used randomization in blocks of four to balance seasonal trends in UV exposure. All participants received $50 per visit (up to $150 across the study), and participants receiving a dosimeter were encouraged to wear it every day and received a compliance payment of $20 per visit if their dosimeter recorded UV at least two days per week (up to $40 across the study). The compliance payments was designed to encourage participants to wear the dosimeter at least during the weekend. Participants were enrolled from April to July 2018 and had two follow-up visits at 3-month intervals. The final visits ran from November to January 2019. All participants from both groups were examined by the same dermatologist who was blinded to their group assignment.

### Wearable UV dosimeters

The sponsor provided the Shade UV dosimeters[5] and a companion smartphone application. The dosimeters were designed to measure the UV index (UVI), a real-time measure of the strength of UV relevant to skin health. The dosimeters measured the UVI every second and aggregated the cumulative dose every 6 minutes. They were designed to be worn on the chest using a magnetic attachment (Figure 1). The sponsor developed an application for both Apple and Android smartphones connected to the UV dosimeter via Bluetooth. While connected to a Shade UV dosimeter, the smartphone application displayed a real-time UV index, real-time cumulative UV exposure, and historical data of daily UV exposure. At enrollment, the device participants were trained to use the dosimeter and select a daily UV dose threshold on the application. This threshold was customizable through the application and could be changed by the participant. Participants’ daily UV exposure would reset to zero at midnight and, as it increases throughout the day, would be compared to the threshold they had set. These UV thresholds were not stored and, therefore, were not available for analysis. Every time the daily UV exposure reaches 20% of the daily UV dose threshold, the app pushes a smartphone notification (e.g., “You have reached 40% of your daily UV dose”). Participants could also inform the smartphone app if they were using sunscreen by indicating the overall SPF but not the body location of the application. The cumulative UV exposure would be divided by the sun protection factor (SPF) during the two hours following sunscreen application before being added to the daily UV exposure.[6]

**Figure 1.**
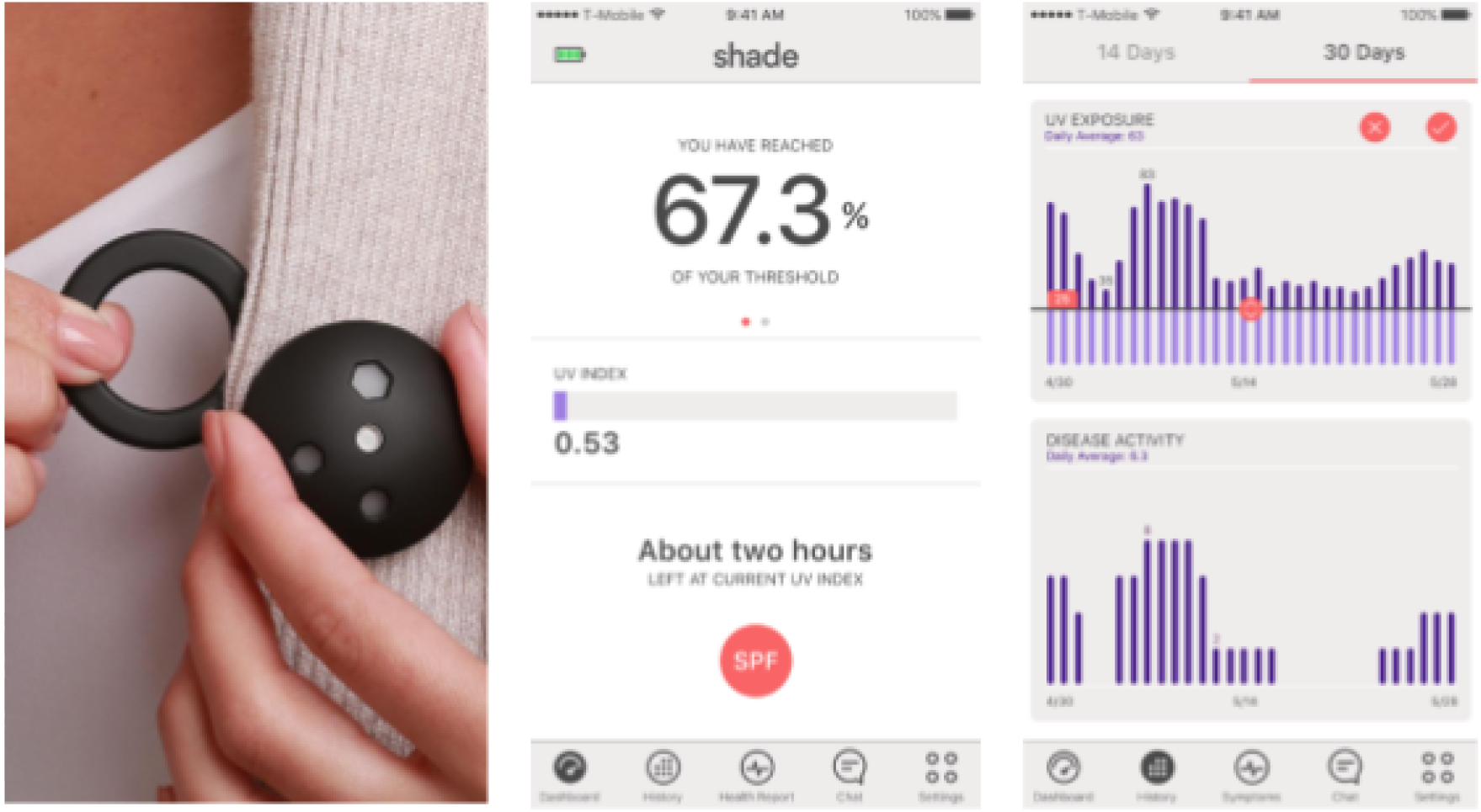
Wearable UV dosimeter, its magnet, and its companion smartphone application.

We note that wearing a UV dosimeter in a specific body area has three inherent limitations: (1) it does not measure UV exposure in other body areas; (2) it does not take into account protective clothing and (3) its measurement depends on its orientation to the sun.

### Safety assessments

Safety assessments included monitoring of adverse events related or possibly related to the device or sun exposure experienced within the study period. Adverse events were to be reported to the clinical principal investigator, the study coordinator, the IRB, and the NCI.

### Efficacy assessments

A single, blinded dermatologist counted AKs and NMSCs on sun-exposed areas (scalp, face, hands) at enrollment and at each subsequent visit (three months after enrollment and six months after enrollment). Pictures of every lesion and its locations were recorded. AKs are defined as keratotic macule(s) or papule(s) on an erythematous base. To ensure that only new AKs or NMSCs after enrollment were counted, each lesion’s location and picture were compared to prior lesions (AK or NMSC). The primary endpoint was the incidence rate of AKs at disenrollment compared to the intermediary visit. Secondary clinical endpoints included the incidence rate of NMSC at disenrollment compared to the intermediary visit. All AKs were treated and eliminated at the time of each visit with cryotherapy, ensuring an accurate calculation of the longitudinal AK incidence. All lesions suspected of being cancerous were biopsied and the diagnoses were confirmed by a blinded pathologist. Other secondary endpoints included scores on three NIH PROMIS 8-question surveys on anxiety, depression, and the ability to participate in social activities.

### Data entry

Case Report Forms (CRFs) were filled out by participants, the study coordinator, and the dermatologist on paper. CRFs were monitored by the sponsor for completeness, consistency, and agreement with underlying medical records periodically during the study. CRFs were scanned and data entered by two people, and discrepancies were reconciled manually.

### Statistical analysis

We first compared all collected clinico-demographic features to identify imbalances between the control and intervention groups. Any feature showing a difference between groups was selected for the subsequent multivariate analyses, regardless of their potential association with AK or cancer. For this covariate selection, we apply a relaxed p-value <0.2 to be more conservative. Our decision to use a threshold p-value of 0.2 is the upper bound of the range recommended for variable selection in linear models.[7]

Incidence rates (IR) of AK and NMSC within 3-month intervals at the intermediary visit and disenrollment were calculated using a multivariate Poisson model that includes clinico-demographic variables with a significant or sub-significant different difference between groups at baseline (p-value < 0.2). All variables are reported in Table 1. The incidence rate ratio (IRR) between groups was calculated using a longitudinal approach, comparing the changes in IRs between the intermediary visit and disenrollment in each group. As the lesions observed at enrollment occurred during an undetermined time interval, the change in IRs was only calculated between the intermediary visit and disenrollment.

**Table 1.**
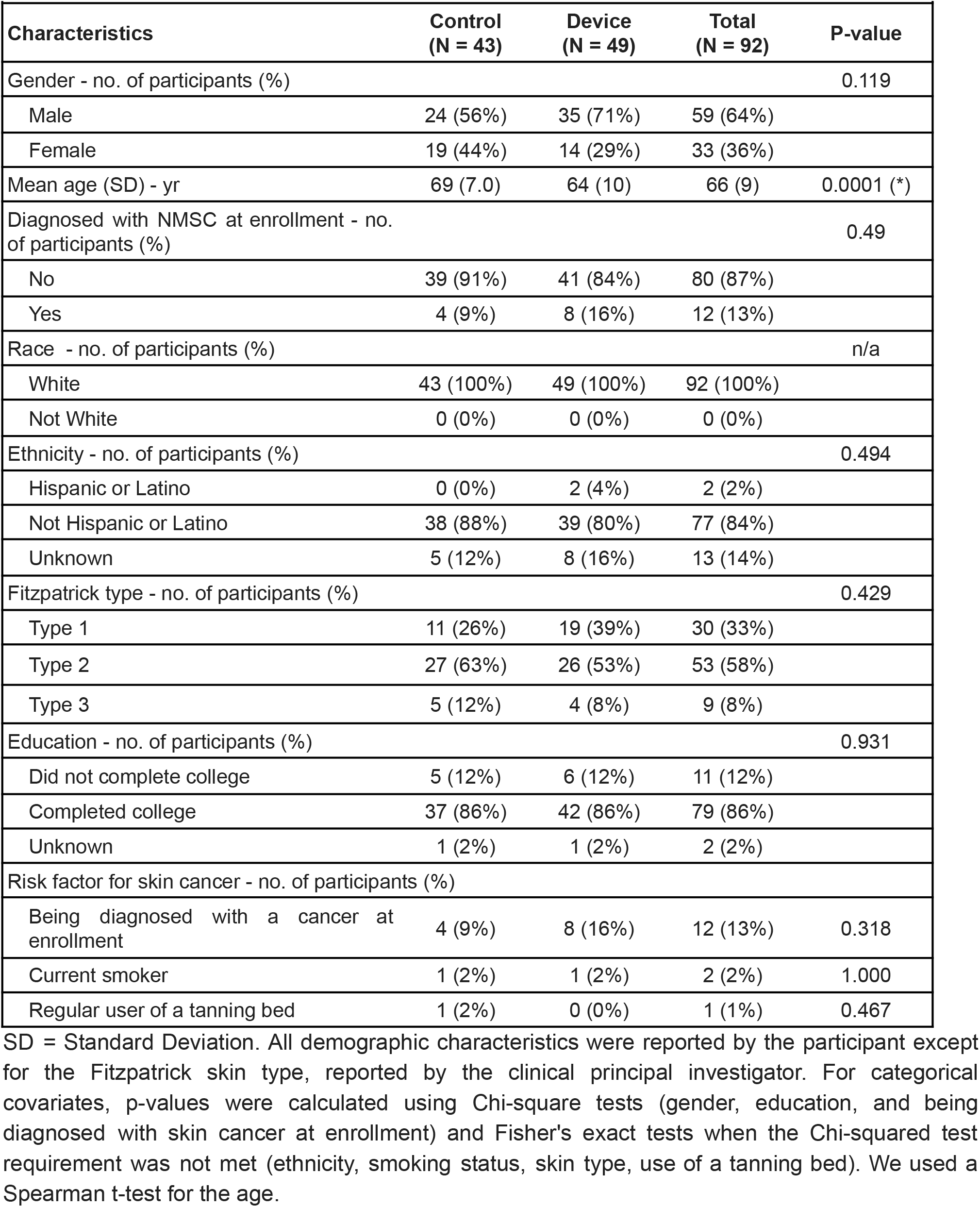
Demographic and clinical characteristics.

**Figure 2. Randomization and Analysis Populations.**
| **Table 1. Demographic and clinical characteristics.** |  |  |  |  |
| --- | --- | --- | --- | --- |
| **Characteristics** | **Control**<br>**(N = 43)** | **Device**<br>**(N = 49)** | **Total**<br>**(N = 92)** | **P-value** |
| Gender - no. of participants (%) |  |  |  | 0.119 |
| Male | 24 (56%) | 35 (71%) | 59 (64%) |  |
| Female | 19 (44%) | 14 (29%) | 33 (36%) |  |
| Mean age (SD) - yr | 69 (7.0) | 64 (10) | 66 (9) | 0.0001 (*) |
| Diagnosed with NMSC at enrollment - no. of participants (%) |  |  |  | 0.49 |
| No | 39 (91%) | 41 (84%) | 80 (87%) |  |
| Yes | 4 (9%) | 8 (16%) | 12 (13%) |  |
| Race - no. of participants (%) |  |  |  | n/a |
| White | 43 (100%) | 49 (100%) | 92 (100%) |  |
| Not White | 0 (0%) | 0 (0%) | 0 (0%) |  |
| Ethnicity - no. of participants (%) |  |  |  | 0.494 |
| Hispanic or Latino | 0 (0%) | 2 (4%) | 2 (2%) |  |
| Not Hispanic or Latino | 38 (88%) | 39 (80%) | 77 (84%) |  |
| Unknown | 5 (12%) | 8 (16%) | 13 (14%) |  |
| Fitzpatrick type - no. of participants (%) |  |  |  | 0.429 |
| Type 1 | 11 (26%) | 19 (39%) | 30 (33%) |  |
| Type 2 | 27 (63%) | 26 (53%) | 53 (58%) |  |
| Type 3 | 5 (12%) | 4 (8%) | 9 (8%) |  |
| Education - no. of participants (%) |  |  |  | 0.931 |
| Did not complete college | 5 (12%) | 6 (12%) | 11 (12%) |  |
| Completed college | 37 (86%) | 42 (86%) | 79 (86%) |  |
| Unknown | 1 (2%) | 1 (2%) | 2 (2%) |  |
| Risk factor for skin cancer - no. of participants (%) |  |  |  |  |
| Being diagnosed with a cancer at enrollment | 4 (9%) | 8 (16%) | 12 (13%) | 0.318 |
| Current smoker | 1 (2%) | 1 (2%) | 2 (2%) | 1.000 |
| Regular user of a tanning bed | 1 (2%) | 0 (0%) | 1 (1%) | 0.467 |

SD = Standard Deviation. All demographic characteristics were reported by the participant except for the Fitzpatrick skin type, reported by the clinical principal investigator. For categorical covariates, p-values were calculated using Chi-square tests (gender, education, and being diagnosed with skin cancer at enrollment) and Fisher's exact tests when the Chi-squared test requirement was not met (ethnicity, smoking status, skin type, use of a tanning bed). We used a Spearman t-test for the age.

The trial was designed for the null hypothesis that the efficacy of the UV dosimeter is less than 25% in reducing the rate of newly-formed AKs over three months. Analyzing over 7,000 patient visits from January 31, 2013 to January 31, 2018 at the department of dermatology at Weill Cornell Medicine, we applied Monte Carlo simulations to determine that a 25% decrease in the number of AKs would be significantly observed (Student’s t-test, p < 0.05; power ≥ 80%) across a population of 102 participants.

## RESULTS

### Trial population

Between April 1, 2018, and July 31, 2018, ninety-seven patients underwent randomization. Fifty were assigned to the device group and received a Shade UV sensor and UV protection counseling. Forty-seven were assigned to the control group and received UV protection counseling only (Figure 2). Skin type, defined by the Fitzpatrick scale (from 1 to 6),[8] was balanced between the device and the control group (Table 1). Gender, skin type, ethnicity, race, education, and known skin cancer risk factors were balanced in the two groups. The mean age of the participants was 66 years. Despite randomization, the participants in the device group were significantly younger than the participants in the control group by five years on average.

**Figure 2.**
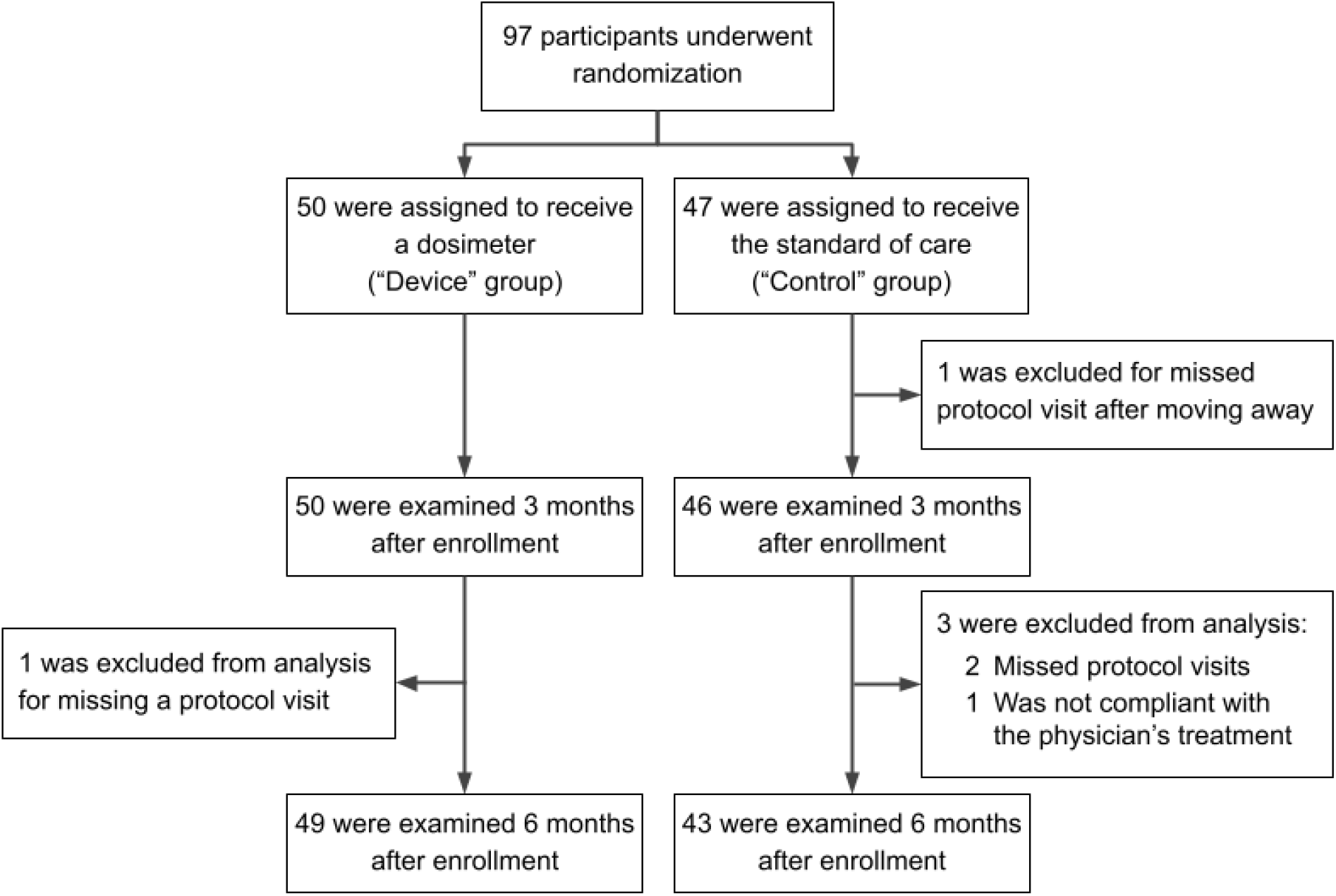
Randomization and Analysis Populations.

### Safety

No adverse events were reported during the trial.

### Efficacy

In Figure 3, we present the incidence rates for AK and NMSC at the intermediary visit (3 months after enrollment) and disenrollment (after summer, six months after enrollment). Six months into the intervention, when comparing the device group to the control group, we measured a non-significant 20% lower ratio of incidence rates of AKs (95% CI = [-41%, 55%], p-value = 0.44) and a significant 95% lower ratio of incidence rates of NMSCs (95% CI = [33%, 99.6%], p-value = 0.024).

**Figure 3.**
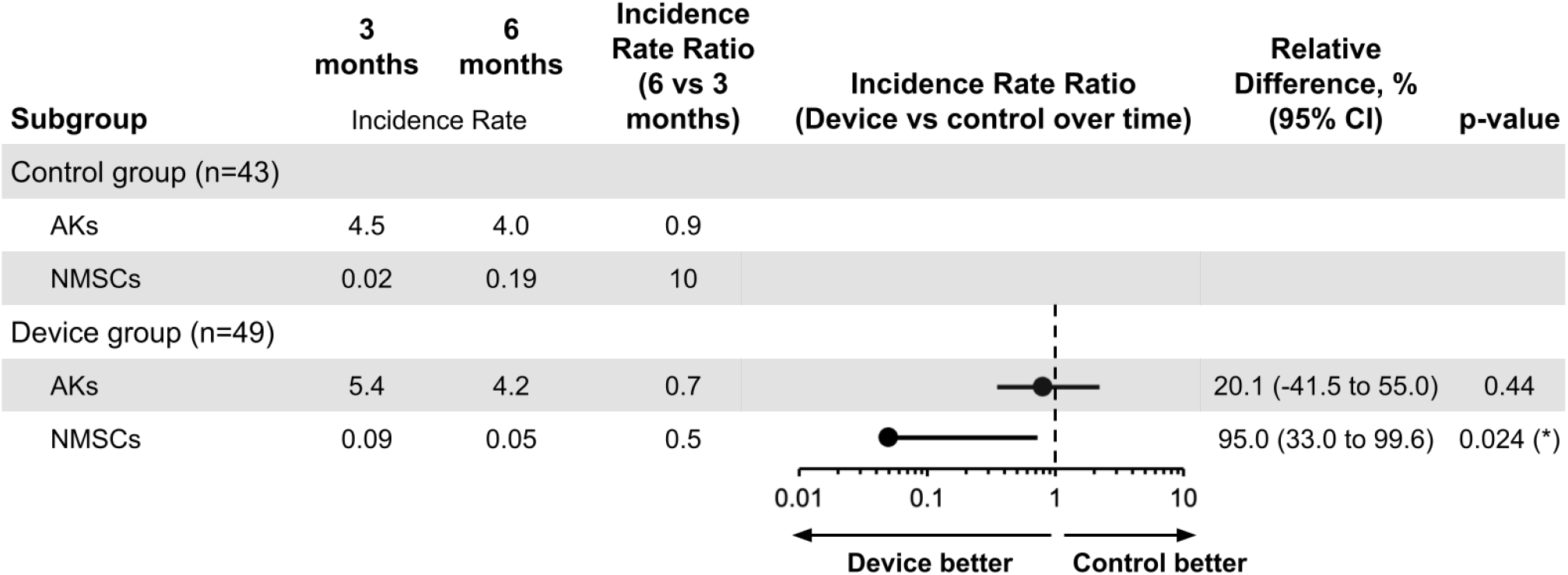
The incidence rate of new AK and NMSC at 3 and 6 months in the intervention. The incidence rate ratio (the ratio of the changes in incidence rates in the two groups), relative differences (1 - IRR, multiplied by 100), and p-values are estimated from a Poisson model that includes all variables whose p-value is below 0.2 as covariates (gender and age). When the covariates gender and age were omitted from the model, the conclusions remained unchanged. The ratio of incidence rates of NMSCs at 6 months was significantly lower in the device group than in the control group (relative difference: 95.0%, p-value = 0.024). This benefit with the device was also observed for AKs but not significantly (relative difference: 20.1%, p-value = 0.44).

Each PROMIS form has 8 questions rated from 1 to 5, for a combined score between 8 and 40. We found a relative decrease of 2.1 points (p-value = 0.010, 95% CI: -3.69, -0.50) in self-reported ability to participate in social events in the device group compared to the control group. We did not find any difference in anxiety or depression.

### UV behavior

Figure 4A displays weekly device compliance and sunscreen use over time. Weekly device compliance is approximated by registering UV once a week. Sunscreen usage was measured by the number of self-reported sunscreen applications through the mobile application. The device compliance remained above 75% for most of the summer and dropped below 50% after November. On average, participants reported applying sunscreen once or twice per week over the summer.

**Figure 4.**
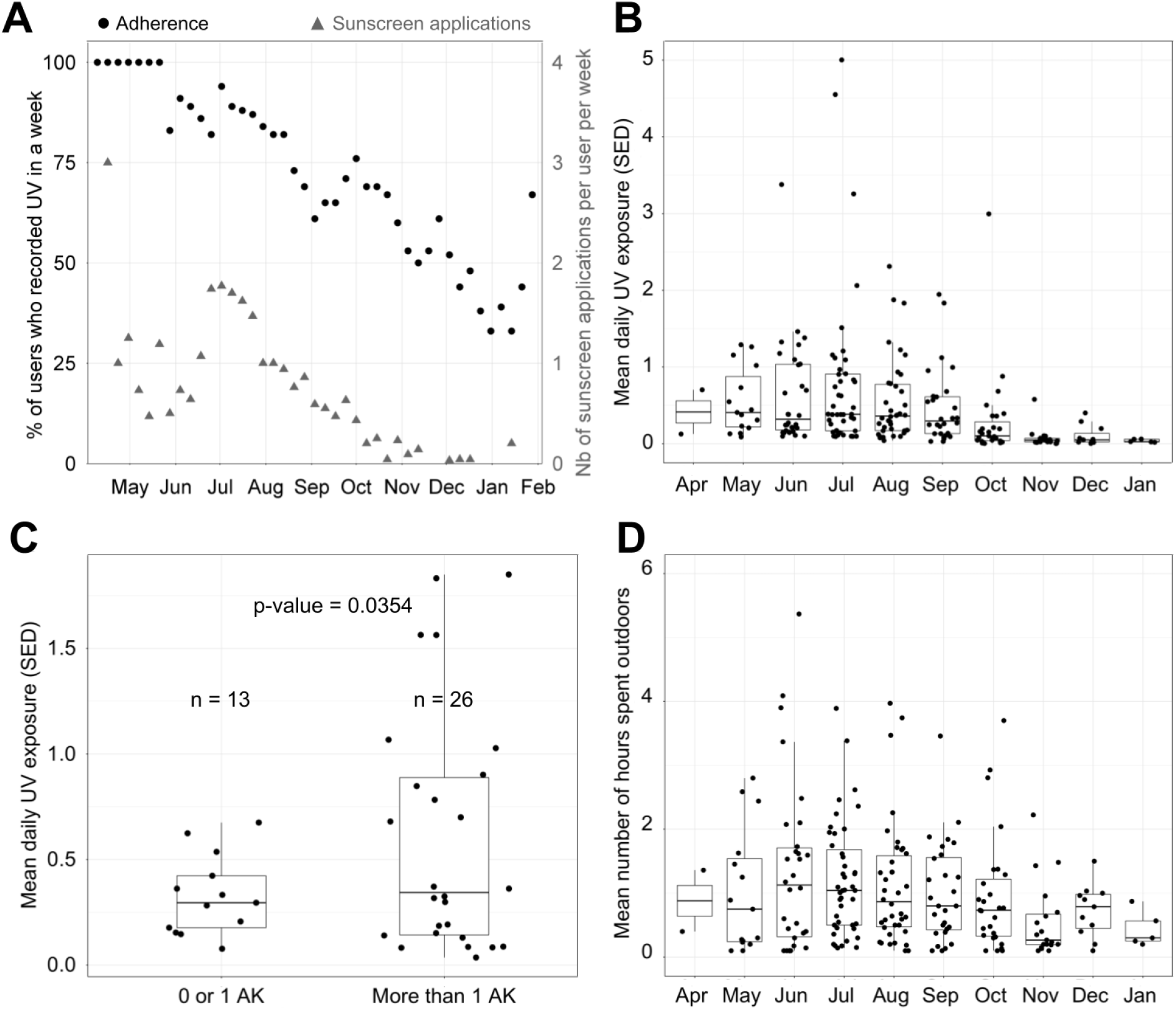
UV exposure data in the device group. A. Percentage of participants who recorded UV exposure in a week and number of sunscreen applications per user per week. B. Distribution of the mean daily UV exposure per month (one data point per participant per month). C. Distribution of the mean daily UV exposure over August and September as a function of the number of AKs measured at disenrollment among participants. Using Welch’s 2-sample t-test, we found that the group with a low number of AKs had a mean UV exposure of 0.33 SED, and the group with a high number of AKs had a mean UV exposure of 0.60 SED (p-value = 0.0354). D. Distribution of the mean daily time spent outdoors per month (one data point per participant per month).

Figures 4B and 4D show the mean daily UV exposure and the mean number of hours spent outdoors per month per participant. Figure 4C clusters participants by the number of AKs diagnosed at disenrollment into two groups and displays the average daily UV exposure over August and September. Device group participants with more than one AK at disenrollment experienced a daily average of 0.60 SED across August and September. In contrast, participants with 0 or 1 lesion experienced a daily average of 0.33 SED across August and September (p=0.0354, Welch’s t-test). This data suggests that sub-erythemal chronic exposure beyond 0.34 SED may contribute to the appearance of lesions. Additional evaluation is needed, as this was not the primary endpoint studied.

## DISCUSSION

This randomized clinical trial was designed to evaluate a novel sun protection strategy over 6 months overlapping one summer where real-time, accurate UV information is provided to participants against the standard of care in UV education. It was powered to detect a 25% reduction in the incidence rate ratio of newly-formed AKs. Our study was underpowered (92 completed the study vs. 102 participants to reach power). As a result, we observed a non-significant 20% lower ratio of incidence rates of AKs (CI = [-41%, 55%], p-value = 0.44) in the device group compared to the control group. However, Figure 4C shows that, in the device group, participants with more than two AKs at disenrollment had a significantly higher average daily UV exposure than participants with less than one AK diagnosed at disenrollment (p = 0.035). This suggests that reducing UV exposure had a significant impact on AK formation.

Additionally, we measured a statistically significant 95% lower ratio of incidence rates of NMSCs (p-value = 0.024, 95% CI: [33%, 99.6%]) after controlling for all variables whose p-value was below 0.2 (age and gender). While these findings suggest that self-managed UV exposure using real-time and personalized UV information might be useful to prevent UV-related skin cancerous lesions, the strong reduction of NMSC incidence rate after a change of UV exposure over only six months is surprising at first. Although we cannot rule out that the observed incidence rate ratio of NMSCs might occur by chance through random sampling, it is unlikely as the p-value of 0.024 indicates that there is only one chance in forty that our observation is a false positive. Also, this result is consistent with the current cancer biology and epidemiology knowledge. If carcinogenesis arises from the accumulation of driver mutations over years or decades,[9] this process is generally described as exponential with age in humans and other mammals.[10] As a result, carcinogenesis has been well explained by the percolation theory, which models human tissue as a network of elements whose probability of transitioning from non-cancer to cancer follows a sudden and dramatic increase.[11] Like other cancers, the NMSC incidence rate increases exponentially with age. [12] This suggests that additional UV-induced driver mutations are exponentially more likely to lead to skin cancer as people age. Importantly, our trial is not the first to measure an intervention’s impact on skin carcinogenesis over a short period. In 2015, Chen et al. demonstrated through a randomized clinical trial that nicotinamide significantly reduces the incidence rate of NMSC over just 12 months in an elderly population.[13] Also, our result is consistent with two established molecular mechanisms. The first one is related to the biology of p53 immunopositive epidermal keratinocytes, also called p53 “patches.” These p53 patches follow UV exposure[14] and are associated with skin carcinoma, with 50% of all skin cancers expressing these mutations.[15, 16] The prevalence of p53 patches increases with age until saturation when people reach the age of 60 years old. [17] Using a murine model, Rebel et al. showed that squamous cell carcinomas (SCC) start appearing after p53 patch saturation, and their count grows exponentially with time when mice continue to be exposed to daily UV.[18] These data also suggest that UV exposure following p53 patch saturation when people are old may accelerate NMSC pathogenesis. Furthermore, this p53 saturation is consistent with the percolation model described by Shin et al. for colorectal tumorigenesis,[11] where the percolation critical transition for skin cancer would occur when the skin is saturated with p53 patches. In addition, UV radiation induces immunosuppression, which in turn triggers a rapid development of NMSC.[19] UV radiation can induce immunosuppression via various mechanisms, including direct immune cell activation and the activation of suppressor immune cells.[20] Both UVA and UVB have distinct effects on immune cell function, and it remains possible that seasonal reduction in UV exposure in our device group may have allowed for increased immune surveillance and NMSC clearance. Together, these biological mechanisms provide a possible rationale for the deceleration of NMSC development in an elderly population following UV avoidance, even after a few months. Furthermore, the two NMSCs reported at disenrollment in the intervention group were diagnosed in participants who stopped using the device a few days after enrollment, providing additional evidence of the impact of the wearable UV dosimeter on NMSC incidence. Finally, given the low number of NMSC measured during the trial, the 95% confidence interval of the incidence rate ratio ranges from 33% to 99%, so we believe a larger trial would show an impact of the dosimeter on the NMSC incidence rate ratio closer to the effect size measured for AKs.

When comparing the device group to the control group, we measured a difference in participants’ self-perceived ability to participate in social activities. The change in scores between enrollment and disenrollment differed by 2.1 points (p-value = 0.01, 95% CI: [3.67, 0.48]) between groups. The control group’s score significantly increased by 1.2 (p-value = 0.04), while in the device group, the score non-significantly decreased by 0.9 (p-value = 0.1). This result suggests that real-time UV data and feedback increase participants’ awareness of UV, while UV counseling leads participants to become overconfident. We did not observe changes, or between-group differences, in self-reported anxiety and depression surveys, suggesting that the device and app do not significantly impact participants’ quality of life.

There are several limitations to this study. First, the number of NMSCs is low, so we could not perform stratified analyses by SCC and BCC, as such analyses would have been underpowered. However, the breakdown of NMSC by type and body location is available in the supplementary information. Also, the study population came from a single recruiting site in New York City, where the highest UVI ranges from 6 and 9 during the summer. It is likely that the impact of the device would vary depending on the UV of the recruiting sites, with a lower impact in low UV versus high UV regions, such as Australia, where similar studies were conducted. While this hypothesis needs to be confirmed in a multicentric study, our study provides a baseline estimate of the efficiency of a wearable dosimeter as a preventive tool in regions with similar UV exposure. In addition, over 85% of our participants completed college, twice the national average; although this could limit the generalization of our findings to the US population, we did not observe any significant impact of education on the impact of the device as the unadjusted incidence rates of NMSC at disenrollment in the device and control groups stratified by education. Besides, the population was followed for one summer only, leaving the possibility that the device’s impact would be short-lived. UV dosimeters, like sunscreen, are seasonal tools mostly used when UV is at the highest, as shown by Figure 4A; therefore, we expect to see an uptake in usage in the Spring. Another limitation to our trial is that we do not know the UV exposure behavior in the control group, so we cannot establish that the device participants have lower UV exposure than the control participants. We chose not to survey control participants’ UV exposure based on their recollection because these surveys are unreliable.[21, 22] We also did not give UV dosimeters to control participants because we wanted a clean comparison to standard-of-care, and we were concerned that simply wearing the device could influence control patients’ UV behavior.[23] Further, wearable UV dosimeters used in this trial measure UV exposure from a single location (the trunk), which is not necessarily representative of sun-exposed body locations (e.g., hands, face, or scalp). Selecting the trunk was a compromise between the ideal location (the forehead, using a headband) and the most suitable location (the wrist, using a watch). The first one would be stigmatizing and could hinder enrollment and observance. The second one is subject to higher within-subject variability and higher variability between subjects than the trunk. Even though a dosimeter on the trunk underestimates UV exposure on the face,[24] this underestimation is true for all participants, and the measurement is stable across participants, thus unlikely to bias our results.

Importantly, device uptake, adherence, and usage measured in this trial do not reflect real-world data. Device participants did not have to purchase the UV dosimeter; they were compensated ($40 over six months) if they wore the device at least two days per week, and broken or lost devices were replaced at no cost to participants. However, wearable UV dosimeters could reach a wide audience in several different ways. They could be part of smartwatches, which upgrade their sensor offerings every other year while keeping their prices stable. Also, if the results of this trial were confirmed through a larger and longer multi-center study, health insurance companies could subsidize the price of wearable UV dosimeters using their savings on skin cancer treatment.[25, 26] Wearable UV dosimeters are not meant to replace established sun protection strategies (clothing, sunscreen, or avoidance). Instead, they would increase the options consumers and healthcare providers have for skin cancer prevention.

## CONCLUSIONS

Over the past few years, consumers have learned about their health by using sensors in wearable devices such as smartwatches. This clinical trial is the first to test the use of an accurate wearable UV dosimeter in the context of skin cancer prevention for an elderly population. The clinical benefit measured in this pilot study shows potential for elderly patients and warrants further investigation through larger and longer studies. Similar studies could be conducted in a younger population focusing on sunburn avoidance. Eventually, such accurate sensors could be integrated into current consumer-grade devices (e.g., smartwatches) as a data-driven, chemical-free sun protection tool. Finally, our trial and Chen’s on nicotinamide[13] suggest that late-stage events play an essential role in skin cancer pathogenesis. Further work is needed to delineate these molecular mechanisms.

## Data Availability

All data produced in the present study are available upon reasonable request to the authors.

## LIST OF ABBREVIATIONS

AK: Actinic Keratosis
BCC: Basal Cell Carcinoma
CI: Confidence Interval
CRF: Case Report Form
IR: Incidence Rate
IRB: Institutional Review Board
IRR: Incidence Rate Ratio
NCI: National Cancer Institute
NIH: National Institute of Healths
NMSC: Non-Melanoma Skin Cancer
PROMIS: Patient-Reported Outcomes Measurement Information System
SCC: Squamous Cell Carcinoma
SD: Standard Deviation
SED: Standard Erythema Dose
UV: Ultraviolet or Ultraviolet Radiation
UVI: UV Index

## DECLARATIONS

### Ethics approval and consent to participate

Consent for the publication of recognizable patient photographs or other identifiable material was obtained by the authors and included at the time of article submission to the journal stating that all patients gave consent with the understanding that this information may be publicly available.

The trial was conducted under the oversight of the Institutional Review Board (IRB) of Weill Cornell Medicine and the National Cancer Institute (NCI). It adhered to applicable guidelines and governmental regulations. The IRB and the NCI approved the protocol and the consent forms. All participants provided written informed consent before enrollment.

### Consent for publication

n/a

### Authors’ contributions

GV was the clinical principal investigator of the study and examined all patients. MB was the clinical study coordinator of the study. PDK was Shade’s clinical study manager. JHZ, CD, PDK, and ELPD designed the clinical trial. PDK, SB, and ELPD digitized the CRFs and prepared the UV exposure data. ELPD and CD did the statistical analysis. KE provided guidance on the data analysis and manuscript preparation. ELPD wrote the manuscript. All authors reviewed and provided input on the manuscript. All authors had full access to all the data in the study and had final responsibility for the decision to submit for publication.

### Competing interests

PDK and ELPD are employees and shareholders of Shade, a startup manufacturing wearable UV sensors and the sponsor of this clinical trial. SB and JHZ are advisors and shareholders of Shade.

### Funding

This work was funded by the National Cancer Institute (contract HHSN261201700005C).

### Availability of data and materials

The datasets generated and/or analysed during the current study are not publicly available due identifiable information present on the case report forms but are available from the corresponding author on reasonable request.

## Acknowledgements

n/a

## Notes

### Clinical Trial

NCT03315286

### Author Declarations

IRB of Weill-Cornell Medicine gave ethical approval for this work

### Summary of Updates

- clarified the discussion - added another author and re-arranged author list

